# Frequent Presenters to Southern New Zealand Emergency Departments differ from other ED attenders

**DOI:** 10.1101/2025.02.26.25322806

**Authors:** Yuta Tamberg, Melyssa Roy, Waldir Rodrigues de Souza, John Eastwood

## Abstract

**Objective:** To compare patterns and characteristics of emergency department visits initiated by frequent presenters and other ED attenders.

**Methods:** We performed a quantitative retrospective data analysis of routinely collected ED data across three public hospitals within New Zealand’s Southern district. The study population comprised: (1) frequent presenters (i.e., patients who visited any of the EDs on 10 or more occasions in any continuous 365-day period between 01/01/2018 and 31/12/2022) and (2) all other ED presenters during the same period. We compared visit-based and per-person metrics characterising presentation flow, utilization patterns and demographics.

**Results:** Frequent presenters constituted 0.6% of all ED attenders (1259 out of 196136) and initiated 6.8% of all ED visits (32541 out of 479975). Visits by frequent presenters showed no seasonality or day-of-the-week patterns and occurred mostly during daylight hours. Frequent presenters differed from non-frequent presenters by having more clustered visits, shorter intervals between presentations, higher presentation severity (more visits were triaged urgent and above), more arrivals by ambulance and police, higher incomplete treatment rate, more referrals and short stay admissions.

**Conclusion:** Frequent presenters make a sizeable contribution to ED load in terms of visit numbers and urgency. The nature of differences between frequent and non-frequent presenters indicate greater and more complex health needs in the former cohort. At the same time the diversity of visits in the frequent presenters’ histories shows potential for redirecting some patients and/or presentations to other care providers.

**Key findings:**

- Frequent presenters make a small portion of total patients, but contribute disproportionately to ED load. High visit numbers and greater acuity indicate more complex health needs in this cohort.
- Frequent presenters differ from other ED attenders by having more clustered visits, shorter intervals between presentations, higher presentation acuity (more visits triaged urgent and above), more arrivals by ambulance and police, higher incomplete treatment rate, and required more referrals and short stay admissions.
- Visits by frequent presenters showed no seasonality or day-of-the-week patterns and occurred mostly during daylight hours.

## 1. Introduction

Overall ED utilisation is increasing faster than population growth, likely due to complex health needs, population ageing, lack of social support, and barriers to accessing primary care among other factors ^1,2^. The same is true for frequent presenters – a group of patients with a history of recurring ED visits ^3^. Frequent presentation is widespread, with FP cohorts found in different countries and healthcare systems ^4^, where they make a heterogeneous group with diverse health needs and sometimes contradictory traits:

- Although FPs generally suffer higher mortality compared to other ED attendees ^5^, a subgroup of ‘chronic’ FPs visits at an undiminishing rate for years ^6,7^.
- Medical care in an ED is deemed appropriate for some FP visits ^8^, while others could be treated by alternative care providers ^9^.
- Finally, FPs are described as either heavy users of many elements of the health system and community care ^10,11^, or as minimally engaged ^12^.
- Understanding the needs and characteristics of local subgroups of FPs is necessary to identify cases likely to benefit from engagement with other care providers. Alternative ways of providing health care to this group may be more beneficial to both FPs and EDs compared to the status quo ^13^.

## 2. Methods

### 2.1. Design and setting

This quantitative retrospective observational study compared cohorts of ED attenders using routinely collected data from three public hospitals within the same District Health Board. The EDs serve areas with different population structures and health needs: Dunedin (153 000 resident population, including ~24 000 students), Queenstown (46 000 resident population, a million tourists annually) and Invercargill (69 000 resident population).

### 2.2. Data sources and study population

The dataset, comprising all visits to Southern District EDs from 01/01/2018 to 31/12/2022, was extracted from the district-wide database, which holds health and administrative data for each hospital. Unique patients were identified by their National Health Index (NHI) numbers. A rolling 365-day sum of ED visits was calculated for each individual. Those who attended 10 or more times at least once in a 12-month interval were deemed ‘broadly-defined’ frequent presenters (FPs) and their entire presentation history was considered as such. All other patients, some of whom may have attended ten times or more in the study period, but not in a continuous 365-day window, were regarded as non-frequent presenters (NFPs). A separate group of ‘narrowly-defined’ frequent presenters comprised individuals who attended 10 times or more in a calendar year. This group was used for numerical comparisons with other studies. Only visits with valid NHIs and date and time of arrival were included.

### 2.3. Variables and outcomes

For each visit, we obtained hospital attended, presentation date and time, mode of arrival, triage and discharge status, presenter’s unique identifier, territorial authority of residence, gender, ethnicity and age at presentation. From a patient’s presentation history, we calculated person-specific measures: visit counts, intervals between visits, temporal clustering index, peak per-year presentation count, acuity index, admission rate and incomplete treatment rate (further details on variables, outcomes and statistical analyses are given in Appendix 1). FPs were compared with non-FPs with respect to: (1) presentation flow, (2) utilisation patterns and (3) demographic characteristics.

#### 2.3.1. Presentation flow

Patient and presentation counts were grouped by calendar years and EDs attended, and a peak per-year presentation count obtained for each individual. Month-by-month, day-of-the-week and hour of the day presentation patterns were compared with chi-squared tests. Data from 2020 were excluded from all temporal analyses to remove COVID-19 lockdown effects ^14^. Intervals between consecutive presentations were measured as days between calendar dates of attendance. Per-person temporal clustering index (tCI) was calculated as described by Stanford et al. ^15^ and compared in FP and NFP cohorts using Wilcoxon test.

#### 2.3.2. Utilisation patterns

We compared the distribution of triage status, mode of arrival and presentation outcomes for FP and NFP visits using chi-squared tests. To account for variability of ED events in a patient’s presentation history, we also calculated FP and NFP per-person acuity index, admission and incomplete treatment rates, and compared them using Wilcoxon or Brown-Mood median tests.

#### 2.3.3. Presenter demographics

FPs and NFPs were described by age, gender and ethnicity. Age-specific attendance rates were calculated as a fraction of locally resident FPs out of the usually resident population of the ED catchment (i.e., its territorial authority).

All statistical analyses were done using R 4.3.1 ^16^. For all tests, threshold significance level was set at 0.01 and multiple comparison corrections applied as needed.

### 2.4. Ethics

Ethical approval and site-specific authorisations were granted by Health and Disability Ethics Committee (ID 18509) and Health Research South.

## 3. Results

### 3.1. Presentation counts

Between 1 January 2018 and 31 December 2022, broadly-defined frequent presenters accounted for 0.6% of all unique presenters and initiated 6.8% of all ED visits. Not all FPs maintained high presentation frequency over multiple years (Table 1, compare broadly and narrowly defined FPs). Per-person visit counts and peak-per-year presentations by NFPs have a log-normal distribution, i.e. strongly right-skewed, with the greatest proportion (52%) attending only once in the entire study period and accounting for 21% of all ED visits. In FPs the pattern is similar, but less extreme (Table 2).

**Table 1.** ED visits for broadly-defined FP, narrowly-defined FP (current calendar year only) and total ED attendance by calendar year and hospital.

| Calendar year | Frequent presenters<br>(narrowly-defined) |  | Frequent presenters<br>(broadly-defined) |  | Total presentations |  |
| --- | --- | --- | --- | --- | --- | --- |
|  | Presenters | ED visits | Presenters | ED visits | Presenters | ED visits |
| 2018 | 236 (0.4%) | 3666 (3.8%) | 910 (1.5%) | 6289 (6.6%) | 60725 | 95938 |
| 2019 | 247 (0.4%) | 3787 (3.8%) | 992 (1.6%) | 7036 (7.0%) | 63423 | 100213 |
| 2020 (COVID first wave) | 216 (0.4%) | 3274 (3.6%) | 984 (1.7%) | 6562 (7.2%) | 58198 | 90883 |
| 2021 | 252 (0.4%) | 3865 (4.0%) | 981 (1.6%) | 6998 (7.2%) | 61302 | 96744 |
| 2022 (COVID second wave) | 190 (0.3%) | 2994 (3.1%) | 867 (1.4%) | 5656 (5.9%) | 62556 | 96197 |
| **Emergency department\*** | Presenters: yearly mean | ED visits: yearly mean | Presenters: yearly mean / 5-year sum | ED visits: yearly mean / 5-year sum | Presenters: yearly mean / 5-year sum | ED visits: yearly mean / 5-year sum |
| Dunedin Hospital | 116 (0.4%) | 1909 (4.2%) | 507 (1.7%) / 758** (0.8%) | 3444 (7.6%) / 17219** (7.6%) | 19582 / 97908** | 45151 / 225755** |
| Lakes District Hospital | 10 (0.1%) | 201 (1.6%) | 63 (0.7%) / 129** (0.3%) | 380 (3.0%) / 1902** (3.0%) | 7549 / 37745** | 12748 / 63740** |
| Southland Hospital | 93 (0.4%) | 1276 (3.3%) | 444 (1.9%) / 655** (0.9%) | 2684 (7.0%) / 13420** (7.0%) | 14019 / 70095** | 38096 / 190480** |
| **Totals** | 846** (0.4%) | 17586** (3.7%) | 1259** (0.6%) | 32541** (6.8%) | 196136** | 479975 |

\* Hospital-specific yearly counts of FPs are given as 5-year averages.
\*\* Hospital specific counts of FPs and total attendance as 5-year totals.

**Table 2.** Temporal patterns in presentations, per-person visit counts and peak per-person per-year presentation counts and intervals between presentations to ED.

|  | Frequent presenters<br>(broadly-defined) | Non-frequent presenters |
| --- | --- | --- |
| **Total presentation counts\*** |  |  |
| Average monthly visits* (± SD) | 541 (± 64) | 7565 (± 387) |
| Average monthly visits in winter [Jun, Jul, Aug]* (± SD) | 539 (± 60) | 7686 (± 485) |
| Average daily visits* (± SD): | 17.8 (± 4.7) | 249 (± 24) |
| Average daily visits* (± SD) to Dunedin Hospital | 9.5 (± 3.2) | 116 (± 14) |
| Average daily visits* (± SD) to Lakes District Hospital | 1.0 (± 1.1) | 34 (± 9) |
| Average daily visits* (± SD) to Southland Hospital | 7.3 (± 3.0) | 98 (± 12) |
| Average weekend daily visits* (± SD) | 17.9 (± 4.6) | 250 (± 23) |
| Average hourly visits* (± SD) | 0.75 (± 0.31) | 10.4 (± 5.45) |
| Maximum hourly visits; interval* | ~1 per hour; 10:00 to 23:00 | ~14-16 per hour; 09:00 to 19:00 |
| Minimum hourly visits; interval* | ~0.3 per hour; 03:00 to 08:00 | ~3 per hour; 02:00 to 07:00 |
| **Per-person presentation counts in 5 years** |  |  |
| 25% | 15 | 1 |
| 50% (median) | 21 | 1 |
| 75% | 28 | 3 |
| 95% | 54 | 7 |
| Max. | 413 | 32 |
| Mean (± SD) | 25.8 (± 23.1) | 2.3 (± 2.2) |
| **Peak per-person per-year presentations** |  |  |
| 25% | 10 | 1 |
| 50% (median) | 12 | 1 |
| 75% | 15 | 2 |
| 95% | 28 | 4 |
| Max. | 163 | 9 |
| Mean (± SD) | 14.8 (± 9.4) | 1.7 (± 1.3) |
| **Repeat visits, %** |  |  |
| same or the next day | 12% | 8% (≥ 2 visits) |
| within 7 days | 33% | 17% (≥ 2 visits) |
| within 28 days | 58% | 29% (≥ 2 visits) |
| **Interval between presentations, days** |  |  |
| Median interval (± MAD) | 17 (± 15) days | 118 (± 113; ≥ 2 visits) |
| Average interval (± SD) | 52 (± 96) days | 242 (± 308; ≥ 2 visits) |
| Average per-person tCI (± SD) | 0.95 (± 0.38) | 0.82 (± 0.58; ≥ 4 visits) |

MAD = median absolute deviation (of the median); tCI = temporal clustering index
\* Temporal metrics of the total visit counts exclude 2020 to remove COVID-19 lockdown effects

### 3.2. Temporal patterns (seasons, days of the week, hours)

The number of ED visits in both FP and NFP cohorts was distributed evenly among summer and winter months, as well as days of the week (Table 2). By contrast, both cohorts had a pronounced hour-by-hour pattern. Fewer visits occurred during the night and early morning hours (02:00 to 08:00; Table 2), although peak FP attendance happened slightly later in the day compared to NFP (χ^2^ p < 0.001).

### 3.3. Intervals and temporal clustering

Among patients who attended ED two or more times, visits were spaced non-uniformly, with a median interval between visits of 17 days for FPs and 118 days for NFPs (Table 2). Over a half (58%) of all visits by FPs and 29% by NFPs occurred within 28 days of the previous arrival. FPs tended to have more clustered presentations (i.e. sets of closely-spaced visits punctuated by longer gaps), compared to NFPs who attended four times or more (Wilcoxon test p < 0.001).

### 3.4. Utilisation patterns

FPs significantly differed from NFPs (all χ^2^ p < 0.001) in their (1) mode of arrival to the ED, (2) urgency of the presentation expressed by triage category, and (3) presentation outcomes (Table 3).

**Table 3.**
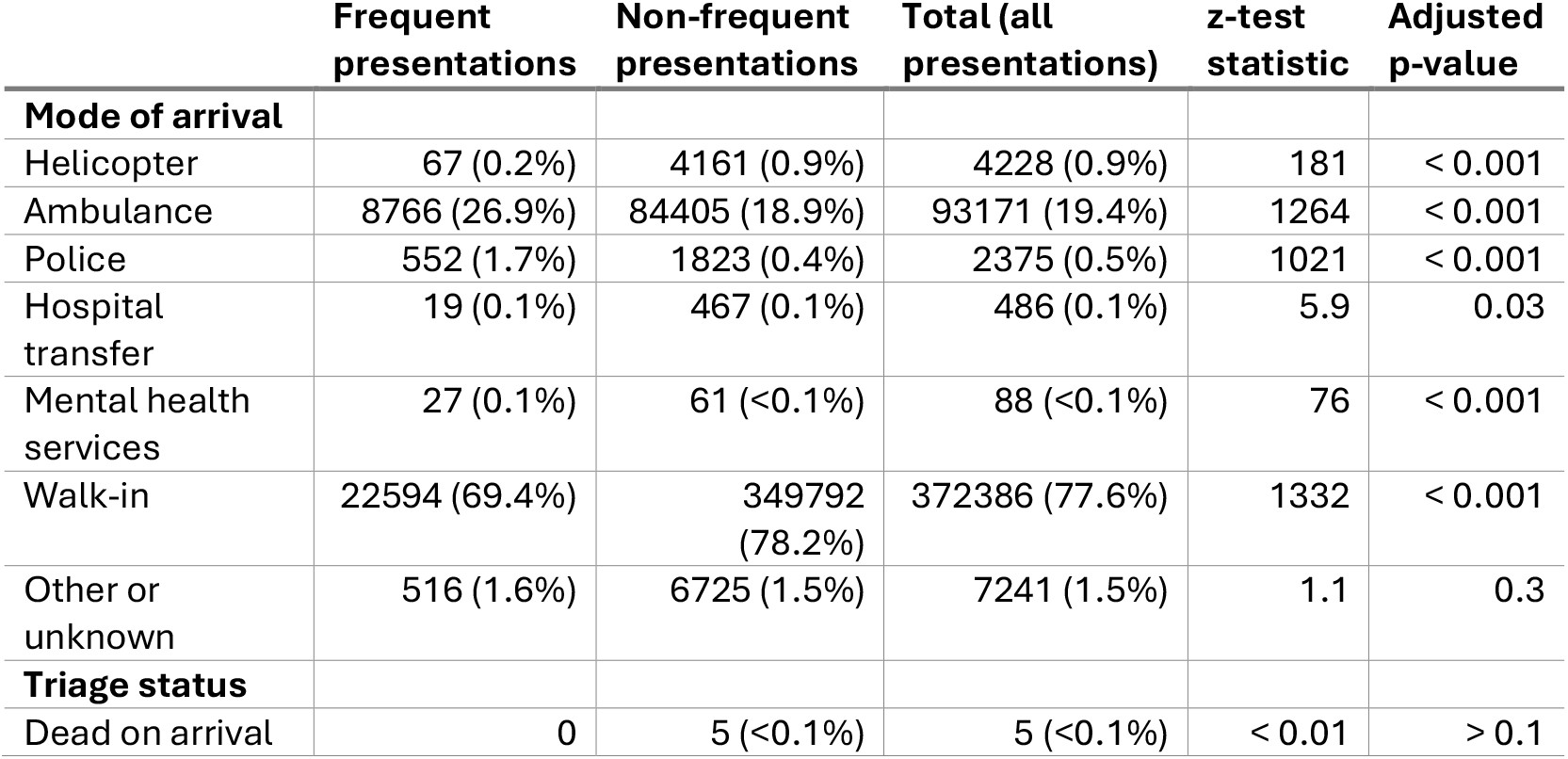
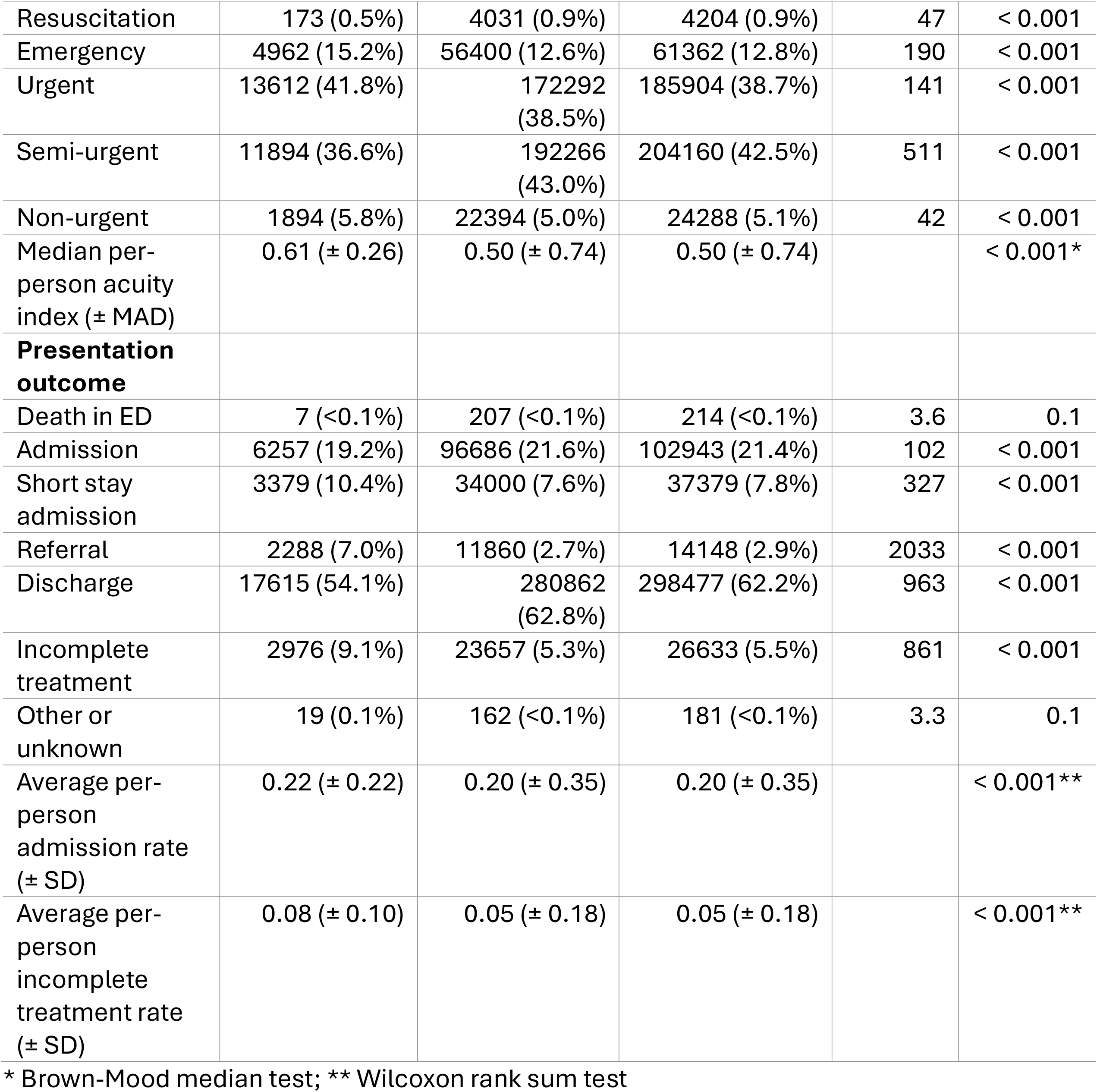
ED visits grouped by mode of arrival, triage status and presentation outcome. Post-hoc row-wise z-test of two proportions, p-values adjusted with Holm method.

For both cohorts, the most typical arrival mode was a ‘walk-in’, although it was less common in FPs (69% vs 78%, z-test p < 0.001). By contrast, FPs had a significantly higher fraction of arrivals by ambulance and police compared to the other attenders (z-tests p < 0.001).

Visits by FPs were most commonly triaged as ‘urgent’, compared to ‘semi-urgent’ in NFPs. In addition, FPs had a lower fraction of ‘non-urgent’ and a higher fraction of ‘emergency’ visits (z-tests p < 0.001) compared to NFP. The distribution of individual acuity levels (fraction of visits triaged as urgent and above) in FPs was log-normal, with a long right tail. In NFPs the distribution was bimodal, dominated by people who only had one visit rated either above or below the urgency threshold. The difference between cohorts was significant (median test p < 0.001).

Treatment was completed in 94% of all ED presentations and discharge home was the most common specific outcome, although less frequent for visits by FPs compared to NFPs (54% *vs* 63%). Visits by the FP cohort more often resulted in a referral, incomplete treatment and a short stay admission compared to NFP visits (z-tests p < 0.001). Hospital admission was less common among FP visits, although the average individual admission rate, i.e., the fraction of patients’ visits resulting in admission, was higher in FPs (Wilcoxon test p < 0.001). Incomplete treatments (i.e., visits aborted when the patient did not wait, left before being seen, refused treatment, or left before treatment completion) were uncommon for all patients, although a larger fraction of FPs had at least one incomplete visit, and the average incomplete treatment rate was also higher in this group (0.08 *vs* 0.05, Wilcoxon test p < 0.001).

### 3.5. Presenter demographics

A lower proportion of FPs were male (44%) compared to NFPs (51%). The median age of FPs was 39 years, compared to 34 years in NFPs. Age at first presentation was distributed unevenly, with distinct peaks in the first year of life and at 15-25 years (Figure 1a, b). Starting at 65 years, age-specific attendance rates per 10 000 population tended to increase in all studied hospitals (Figure 1c). Other demographic parameters are given in the Appendix 2.

**Figure 1.**
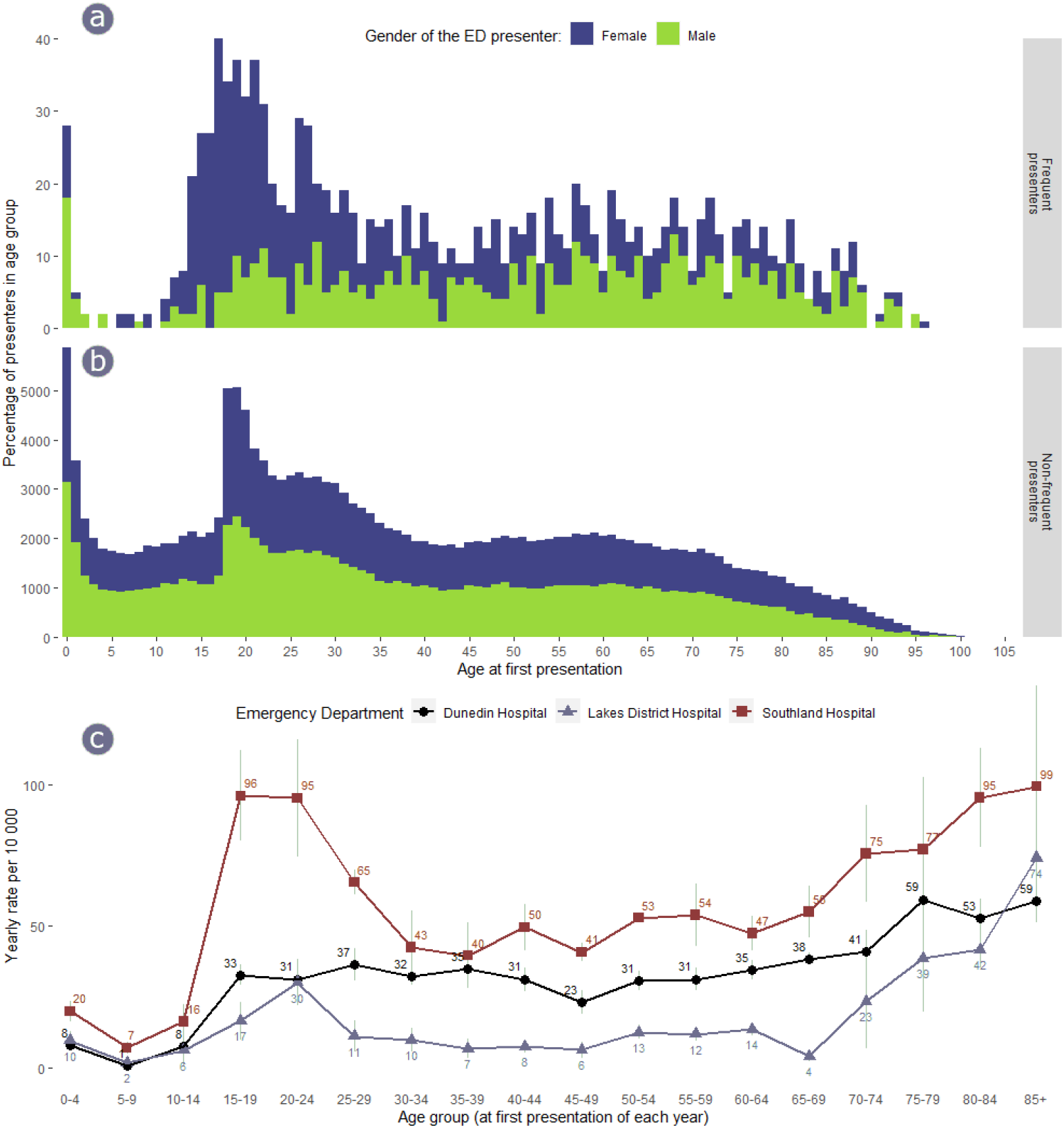
a, b. Age at first presentation in FP and NFP cohorts. c. Age-specific rates of resident FP per 10 000 resident population of the corresponding ED catchments.

## 4. Discussion

The threshold defining frequent presenters usually ranges from 4 to 12 visits in 12 months ^13^. In publications using the same threshold as this study (10+ visits), frequent presenters often constitute 0.2-0.4% of all ED attenders and account for 1.1-4.6% of visits (Table 4). In our study FPs represent a higher fraction of ED attenders and visits (0.6% and 6.8%), partially due to our broader definition: all individuals who attained FP status at least once are considered as such throughout the study (see also Korczak et al. ^17^). The yearly FP attrition rate is 60-80% ^6,7,18,19^, i.e., only 20-40% of one year’s FPs remain so in the following year(s). They often continue to visit EDs, but at rates below the threshold. Thus, combining their entire presentation histories over the study period accounts for a higher than previously reported fraction of total attendance.

**Table 4.** Frequent presenter characteristics. Negative values in bold. CY – calendar year, FY – financial year.

| Source<br>Author, year | Definition,<br>visits per<br>year | % FP<br>patient<br>s (10+<br>only) | % FP<br>visits<br>(10+<br>only) | Average/m<br>edian age,<br>FP | % male<br>FP | % urgent visits<br>by FP (FP –<br>NFP, %) ** | % arrivals by<br>ambulance by<br>FP (FP – NFP, %)<br>** | % admissions<br>by FP (FP –<br>NFP, %) ** | % of incomplete<br>treatment visits<br>by FP (FP – NFP,<br>%) | Year | Country |
| --- | --- | --- | --- | --- | --- | --- | --- | --- | --- | --- | --- |
| Moore et al., 2009<sup>25</sup> | ≥ 4 / ≥ 10<br>(all) | 0.2% | 2.3% | 45.6 (av) | 69.5% | 54.3 (15.5%) | 24.8 (-) | 20.6 (-) | 4.1 (-) | FY 2006/2007 | UK |
| Dent et al., 2010<sup>18</sup> | ≥ 10<br>(adults) | - | 1.9% | 45 (av) | 64% | - | 60 (-) | 39 (17%) | 12 (-) | TY 2007/2008 | UK |
| Kennedy &<br>Ardagh, 2004<sup>6</sup> | ≥ 10<br>(adults) | - | 1.7% | 41 (med) | 57% | - | - | - | - | CY 1997 | NZ |
| Jelinek et al.,<br>2008*<sup>3</sup> | ≥ 10<br>(adults) | 0.4% | 3.7% | 49 (av) | 56% | 42 (14.1%) | 27.9 **(-1.1%)** | 11.5 (6.8%) | 11.5 (6.8%) | CY 2001-2006 | Australia |
| Stanford et al,<br>2024<sup>15</sup> | ≥ 10 (all) | - | 4.6% | - | 59.5% | - | - | 34 (-) | 7 (-) | FY 2015-2021 | Australia |
| Salazar et al.,<br>2005<sup>27</sup> | ≥ 10<br>(adults) | - | 1.1% | 55 (av) | 67.4% | - | - | 8.3 **(-1.8%)** | - | CY 2000 | Spain |
| This study<br>(broadly defined<br>FP) | ≥ 10 (all) | 0.6% | 6.8% | 39 (med) | 44% | 57 (5%) | 27 (8%) | 19 **(-3%)** | 9 (4%) | CY 2018-2022 | NZ |
| This study<br>(narrowly defined<br>FP) | ≥ 10 (all) | 0.4% | 3.7% | 40 (med) | 42% | - | - | - | - | CY 2018-2022 | NZ |
| Mandelberg et al.,<br>2000<sup>19</sup> | ≥ 5 (all) | - | - | - | 70.4% | 54.9 **(-9.6%)** | 25.6 (0.6%) | 12.0 **(-0.3%)** | 13.7 (5.8%) | FY 1993-1998 | USA |
| Markham &<br>Graudins, 2011<sup>28</sup> | ≥ 7<br>(adults) | - | - | 47 (med) | 51.6% | - | 51 (20%) | 31.9 (4.7%) | 10.1 (4.2%) | TY 2009/2010 | Australia |
| Lago et al., 2019<sup>7</sup> | ≥ 7 (all) | - | - | - | 53% | 38.2 (3%) | - | 36.4 (5.6%) | - | FY 2005-2015 | Australia |
| Cordell et al.,<br>2022<sup>29</sup> | ≥ 4 / ≥ 10<br>(adults) | 0.3% | - | 46 (med) | 47.3% | 56.7 (8.5%)** | 30.5 (10.8%)** | - | - | CY 2018 | Australia |
| Zhou et al., 2023<sup>30</sup> | ≥ 4<br>(adults) | - | - | - | 55.1% | 61.0 (-) | 42.1 (-) | - | 5.5 (-) | CY 2019 | Australia |
| Same<sup>30</sup> | same | - | - | - | 46.2% | 90.5 (-) | 46.2 (-) | - | 4.7 | same | Canada |
| Dent et al., 2003 <sup>8</sup> | Top 500 presenters / 10+ | - | 1.6% | 50 (med) | 56.5% | 35.1 **(-12.2%)** | - | 20.1 **(-6.3)** | 4.5 (7.8%) | 64 months 1996-2002 | Australia |
| Korczak et al., 2024 <sup>17</sup> | ≥ 20 (adults) | - | - | - | 55.6% | 39.7 (-) | 34.9 (-) | 18.6 (-) | 11.5 (-) | CY 2015-2020 | Australia |
| Murphy et al, 1999 <sup>26</sup> | ≥ 10 | 0.2% | 2% | - | - | 14.6*** **(-1%)** | - | - | - | CY 1995 | Ireland |

\* FP and NFP rates are re-calculated from raw data reported by the authors for many different attendance groups (1-4 and 4-9 combined as NFP and 10-20, 20-30 and 30+ as FP). Numbers are taken from tables and reconstructed from graphs.
\*\* data for the index visit in presentation history; otherwise aggregated across all visits by the FP/NFP group.
\*\*\* inverse of the percentage of non-emergency visits reported in the article.

If we limit the FP definition to those who qualify only for one calendar year (‘narrowly-defined’ FPs in Tables 1 and 4), the resulting fractions become closer to other published values. This definition also enables direct numerical comparison with a 2004 New Zealand study by Kennedy and Ardagh. The authors reported 72 to 85 FPs a year attending Christchurch hospital ED between 1997 and 2000 ^6^. The Christchurch population in 1997 is estimated at 329,200, which gives a rate of ~2.4 FPs per 10 000 population. We observed 8.9 FPs per 10 000 population in 2018 in Dunedin, a notable increase in FP rates over the last two decades in Southern NZ.

Furthermore, we used a rolling time window instead of static calendar/financial years to identify FPs and counted visits to all three EDs towards the threshold, contributing to higher counts. Another study using a rolling time window ^15^ also reported a higher fraction of FP visits (4.6%). Consequently, some previous studies may have undercounted FPs, suggesting this phenomenon could be more widespread.

Patient re-presentation rates, visit clustering and presentation outcomes can be used to compare the complexity of health care needs among populations. We found that 58% of FP visits occurred within 28 days of previous arrival compared to 29% in NFPs. These values are notably higher than the general per-visit re-presentation rate of 18% reported by Moore et al. ^20^. The median interval between repeat visits was also shorter (5 days) compared to the Australian study (8 days) ^20^. In another study, a subgroup of FPs with mental health, drug, alcohol or social diagnoses had a higher visit clustering index ^15^. Incomplete treatment accounts for 5-7% of all outcomes (^20,21^, present study), and is consistently higher in FPs than NFPs (Table 4). Similarly, referrals accounted for less than 10% of all outcomes, but were twice as common in FPs. These findings indicate FPs have more complex health needs that could benefit from involvement of other care providers.

Triage severity, arrival by ambulance, admission rates and length of stay (not covered) are markers of ED resource utilisation ^22^. In many studies FPs differ from other ED attenders along these metrics (Table 4), although inconsistently. These dissimilarities likely stem from differences between frequency thresholds, study settings and durations, and underlying population heterogeneity. Thus, local information is still needed to describe frequent attenders. Although the proportion of high-acuity visits, arrivals by ambulance and admissions varies between studies (8% to 90%, usually around 30-40%; see Table 4), it never falls close to zero, so the overall impact of FPs on emergency departments is undeniable.

In a 2005 study Bamezai and colleagues found that ~85% of ED costs are workforce expenses ^23^. Since staffing is deployed flexibly according to demand, there is little opportunity for EDs to benefit from economies of scale. In other words, the cost of one additional visit (marginal cost) is close to the average visit cost. Thus, redirecting even some presentations away from the ED would make a difference to the cost of care.

Demand prediction helps reduce ED overcrowding and improve resource allocation. Calendar variables are most commonly used to predict general attendance ^24^. Frequent presenters, however, have no visit seasonality, no preference for day of the week, and higher visit frequency during daylight hours (although slightly later in the day) ^18,20,25,26^. Thus, FP-specific demand prediction does not seem practical. At the individual level, however, temporal clustering may predict near-future visits by a particular patient, opening possibilities for better engagement.

Contrasting views exist on whether ED visits by frequent attenders are amenable for redirection ^8,9^. The average higher acuity of FP visits indicates that ED care is often appropriate for this cohort. Nonetheless, FPs are heterogenous and some individuals may benefit from receiving care from other providers. Additionally, high-acuity visits rarely comprise the entire presentation mix of an individual patient: up to 90% of FP patients have 10% of visits or more triaged semi-urgent or non-urgent and could be managed elsewhere.

## 5. Limitations

The frequency threshold of 10 presentations in a 365-day interval is somewhat arbitrary; other values are also commonly used ^5,7^. Temporal analyses exclude 2020 due to the effects of NZ COVID-19 lockdowns on ED presentations ^14^. Contradictory values sometimes occurred in the demographic variables and addresses. It was often impossible to resolve these data conflicts, so reported demographic parameters and resident rates may contain minor inaccuracies. Length of stay, which is an important descriptor of ED visits, was not considered.

## 6. Conclusions

FPs make up a considerable fraction of total ED attendance. The rate of FPs in ED catchment population has increased in NZ in recent decades. Frequent presenters are heterogenous across years, countries and healthcare systems; thus, local information is still required to understand them. FP presentation patterns do not favour cohort-level demand prediction, however, the diversity of FP presentation histories suggests the possibility of redirecting some patients and/or visits to other care providers. Given their use of ED resources, even a few diverted visits would impact ED expenditure and service delivery.

## 7. Acknowledgements

Authors are grateful to Clementine Trengrove (Data Analyst, Health New Zealand | Te Whatu Ora) and wish to acknowledge Dr. Jim Ross (Senior Lecturer, University of Otago).

## 8. Author contribution

Concept (JE), study design (JE, YT), ethics proposal (all authors), data analysis (YT), manuscript preparation (YT), manuscript review (all authors).

## 9. Competing interests

None declared.

## 10. Data availability statement

The data that support the findings of this study are not publicly available. R scripts are available from corresponding author upon reasonable request.

## Notes

### Competing Interest Statement

The authors have declared no competing interest.

### Funding Statement

This study did not receive any funding

### Author Declarations

Health and Disability Ethics Committee of the Ministry of Health of New Zealand gave ethical approval for this work (ID 18509)

