## Appendix 1 for "Frequent Presenters to Southern New Zealand Emergency Departments differ from other ED attenders"

**Appendix 1: Details of statistical analyses**

### 1.1. Presentation flow characteristics

**Presentation counts:** we calculated overall ED event counts as well as per-person counts for frequent and non-frequent presenters, grouped by calendar years and EDs attended. During the study period, some of the presenters moved in or out of the district, some were born or died. To characterise this shifting population better, we also calculated a peak per year presentation count for each individual (after Stanford et al., 2024). For this, we produced a cumulative sum of visits within a sliding 365-day window and selected the maximum value per person.

**Visit periodicity**: we explored three higher-level temporal patterns of ED events initiated by frequent and non-frequent presenters: (1) month-by-month seasonality, (2) day of the week and (3) hour of the day visit distributions. All presentations from 2020 were excluded to remove the COVID-19 lockdown effects. The rest of ED events were grouped by presentation date at the level of (i) month and year, (ii) day, month and year, and finally (iii) hour, day, month and year, counted within group, and averaged for month, day of the week and hour of the day. Where no visits were recorded for a particular time interval, a backbone was applied. Resulting periodicity distributions within each presenter group were compared to the artificially constructed uniform distributions, which repeat (i) an overall within-group monthly average 12 times, (ii) daily average 7 times and (iii) hourly average 24 times using chi-squared tests. In all cases when the test demonstrated significant differences, a post-hoc analysis of raw residuals was undertaken to find specific deviations contributing most to the test outcome.

**Presentation intervals and temporal clustering:** the raw intervals between consecutive presentations were measured as days between calendar dates of attendance, for frequent and a subset of non-frequent presenters with two or more visits in the study period. To describe the distribution of intervals, we calculated the percentiles of repeat presentations occurring at the following intervals: (1) on the same or the following day, (2) within a week and (3) within four weeks of the original presentation. Central tendencies (mean and median) and variabilities (standard deviations and median absolute deviations respectively) were also calculated.

To estimate per-person temporal clustering index for those who visited ≥4 times, raw intervals were transformed into a logarithmic scale with base 3 and their median absolute deviations were calculated (following Stanford et al., 2024). Low index indicates uniformly spaced presentations, whereas higher values point towards uneven intervals, i.e. some presentations occurring in close succession and separated by longer gaps. The resulting set of personal tCIs was grouped (1) by the total count of visits in the person's presentation history and (2) by frequent and non-frequent presenter status. The former were considered for linear regression, but analysis was impossible due to violation of assumptions. The latter distributions were compared with Wilcoxon test and reported as per-group averages ± SD.

### 1.2. Utilisation pattern characteristics

**Triage, mode of arrival and presentation outcomes**: we compared raw (per-visit) frequencies for these categorical variables using Pearson chi-squared tests of independence to determine differences between frequent and non-frequent presenters. When significant differences were detected, residual analysis (Sharpe, 2019) and row-wise z-tests with Holm correction for multiple comparisons (Kassambara, 2023) were performed to identify the specific variable levels that contribute most to observed differences.

In the individual presentation history, especially for frequent presenters, we often see a combination of ED events with different triage and different outcomes. To capture this variability at a per-person level (instead of per-presentation level), we calculated acuity index as well as admission and incomplete treatment rates for each ED attender. The per-person acuity index is the sum of ED events triaged as urgent, emergency or resuscitation, divided by the total visit count for the individual. Similarly, a per-person admission rate was obtained as a count of ED events resulted in admission, and incomplete treatment rate as a count of aborted ED events (when the presenter left before being seen, refused treatment or self-discharged before treatment was completed), divided by total ED visits. Subgroups of frequent and non-frequent presenters were compared with Wilcoxon or Brown-Mood median tests.

#### 1.3. Presenter demographics

Unlike variables considered above, which change from presentation to presentation, demographic characteristics of a person are usually more stable. The three hospital Emergency Departments examined here differ in size and serve different populations with dissimilar ages, ethnic structures and health needs. Thus, each of the EDs was characterised separately. Age was selected as a key demographic parameter to obtain specific attendance rates. We identified an "ED catchment" as a territorial authority where ED is located (i.e. Dunedin City, Queenstown-Lakes District and Invercargill City) and obtained a portion of frequent presenters resident in the catchment from the usually resident population of the corresponding territorial authority and split into 5-year age groups. Some frequent presenters visited two or three different hospitals during the study period, thus appearing more than once in the ED-specific counts. In addition, among 1253 frequent presenters with a valid street address recorded at least once in EDIS, 671 (54%) have more than one address, and 117 (9%) have five or more different addresses listed. Such address changes sometimes place the same individuals both inside and outside the given ED's catchment area at different times. As a result, they are also counted more than once. No visiting frequent presenter rates were calculated.

### References specific to this Appendix

Kassambara, A. (2023). rstatix: Pipe-Friendly Framework for Basic Statistical Tests. R package version 0.7.2. https://rpkgs.datanovia.com/rstatix/

Sharpe, D. (2019) "Chi-Square Test is Statistically Significant: Now What?". Practical Assessment, Research, and Evaluation: Vol. 20, Article 8. DOI: https://doi.org/10.7275/tbfa-x148
