## Appendix 2 for "Frequent Presenters to Southern New Zealand Emergency Departments differ from other ED attenders"

**Appendix 2:** Median age, gender and ethnic composition of presenters to three studied EDs (5-year total).

| | **Frequent presenters (broadly-defined)** | **All presenters** |
|---|---|---|
| **Dunedin Hospital: total** | 758 (100%) | 97908 (100%) |
| Median age | 40 | 36 |
| Gender | | |
| Female | 426 (56%) | 48407 (49.5%) |
| Male | 332 (44%) | 49492 (50.5%) |
| Other or unknown | - | 9 |
| Ethnicity | | |
| Maori | 77 (10%) | 8312 (9%) |
| Pacific | 18 (2%) | 2968 (3%) |
| Asian | 5 (1%) | 3995 (4%) |
| European and Other | 657 (87%) | 82343 (84%) |
| Unknown | 1 | 290 |
| **Queenstown-Lakes Hospital: total** | 129 (100%) | 37745 (100%) |
| Median age | 38.5 | 30 |
| Gender | | |
| Female | 64 (50%) | 17460 (46%) |
| Male | 65 (50%) | 20268 (54%) |
| Other or unknown | - | 17 |
| Ethnicity | | |
| Maori | 12 (9%) | 1690 (4.5%) |
| Pacific | 4 (3%) | 497 (1%) |
| Asian | 2 (1.5%) | 2962 (8%) |
| European and Other | 110 (85%) | 32281 (85.5%) |
| Unknown | 1 | 315 (1%) |
| **Southland Hospital: total** | 656 (100%) | 70095 (100%) |
| Median age | 38 | 36 |
| Gender | | |
| Female | 377 (58%) | 33815 (48%) |
| Male | 278 (42%) | 36272 (52%) |
| Other or unknown | - | 8 |
| Ethnicity | | |
| Maori | 99 (15%) | 9463 (13%) |
| Pacific | 18 (3%) | 1868 (3%) |
| Asian | 5 (1%) | 2601 (4%) |
| European and Other | 533 (81%) | 55959 (80%) |
| Unknown | - | 204 |
